# Types, Subtypes and Positivity Rates of Seasonal Influenza in Uganda, 2019–2023

**DOI:** 10.64898/2026.08.29.26361662

**Authors:** Martha Annet Nankya, Nicholus Owor, John Timothy Kayiwa, Julius Julian Lutwama, Samuel Gidudu, Hildah Tendo Nansikombi, Gloria Bahizi, Alex Riolexus Ario

## Abstract

**Background:** Seasonal influenza, commonly known as flu, is an acute respiratory, highly contagious illness caused by influenza viruses. A clear understanding of influenza seasonality is crucial for guiding prevention and treatment strategies, including decisions on vaccination timing to prevent outbreaks. While well documented in temperate regions, data on influenza epidemiology in tropical areas, particularly sub-Saharan Africa, remain limited. We described the types, subtypes and positivity rate of seasonal influenza in Uganda during 2019–2023.

**Methods:** This study used descriptive statistics to understand the characteristics of the data, and then employed multivariable logistic regression to identify factors independently associated with a positive influenza test result. The regression analysis examined the impact of sex, sample type, time of year, and year on influenza positivity, quantifying these associations with odds ratios, confidence intervals, and p-values to assess their statistical significance.

**Results:** Of 18035 samples, 938 (5.2%) tested positive for influenza, with a higher prevalence in Influenza-Like Illness (ILI) cases (5.7%) compared to Severe Acute Respiratory Infection (SARI) cases (3.7%). Influenza A was the predominant type (69.7%), followed by influenza B (29.7%, all B. Victoria), with rare co-infections. Among influenza A subtypes, A/H3N2 was most common (65.1%), followed by A/H1N1pdm09 (33.5%). Multivariable logistic regression revealed that the male gender, the month of March, and decreasing year were associated with increased influenza positivity, while SARI sampling and other months showed reduced positivity.

**Conclusion:** The seasonal influenza positivity rates from 2019 to 2023 and the predominance of Influenza A and H3N2 highlight the need for sustained surveillance in Uganda. Given Influenza A’s high genetic variability and potential for novel strain emergence, monitoring circulating strains, informing vaccine development, and implementing targeted interventions for high-risk groups and regions are critical to controlling and preventing outbreaks.

## Background

Seasonal influenza, commonly known as flu, is an acute respiratory, highly contagious illness caused by influenza viruses with effects in the nose, throat, and to some extent lungs. The illness can be graded as either mild or severe; and could result into death. Severity is dependent on factors such as patient’s age, underlying health conditions, immune response and the specific strain of the virus (1). Influenza virus attacks mostly young children, probably due to their lack of immunity from prior exposure to the virus (2,3). Conversely, many adults also contract influenza but may not seek medical attention due to milder symptoms, potentially leading to an underestimation of its true burden in older age groups (4).

Influenza virus affects nearly 10% of the global population each year, resulting in approximately half a million deaths annually worldwide (5). Investigations of seasonal influenza outbreaks in Sub-Saharan Africa in 2021 revealed high mortality rates (2.8-16.5 per 100,000) in Sub-Saharan Africa (6). A study conducted in Uganda revealed that from 2010 to 2015, 11.2% of tested cases of Severe Acute Respiratory Infection (SARI) and Influenza-Like Illness (ILI) were positive for the virus. The burden was higher among patients with ILI (12.8%) compared to those with SARI (8.5%) (7). Over 70% of influenza-associated pneumonia hospitalizations in Uganda occur in children under five years, with the highest rates in those under two years. School-going children aged 5–14 years exhibit the highest influenza positivity rates, and the disease peaks during the rainy season, particularly from June to November. Influenza A is more prevalent than B, and cases are clustered in urban and peri urban areas (8–10).

Influenza surveillance has been a critical public health strategy to monitor and mitigate the impact of the disease in Uganda and globally. Insufficient influenza surveillance infrastructure, testing practices, and healthcare services can worsen the clinical burden of influenza in low and middle-income countries (11). Since 2007, Uganda has implemented a national sentinel surveillance system for influenza, spearheaded by the Uganda Virus Research Institute (UVRI) (12), which serves as a World Health Organization (WHO)-designated National Influenza Center. This system tracks circulating strains, detects novel viruses, and provides data to inform vaccine development and public health interventions. Literature has it that, influenza A, in 2016, was the most predominant virus in Uganda (13). Influenza A is of particular concern due to its pandemic potential (14). Unlike other types, Influenza A undergoes subtyping based on its hemagglutinin (H) and neuraminidase (N) surface proteins, such as H3N2 or H1N1. This virus exhibits high genetic variability through antigenic drift and shift, enabling the emergence of new strains capable of causing widespread epidemics or pandemics (15). Understanding the influenza positivity rate, seasonal patterns, and geographical distribution is crucial for strengthening surveillance, facilitating early epidemic detection, and guiding effective prevention and control efforts (16). We described the types, subtypes and positivity rate of seasonal influenza in Uganda from January 2019 to December 2023.

## Methods

### Study design and setting

We conducted a descriptive analysis of samples that were tested for seasonal influenza in Uganda from January 2019 – December 2023. In Uganda, influenza surveillance operates through a network of hospital based sentinel sites. These sites are selected through purposive sampling, guided by World Health Organization (WHO) recommendations and in collaboration with the Ugandan Ministry of Health. The selection criteria included representation of both rural and urban settings, proximity to trade routes, major landing sites for migratory birds, and live bird markets. Based on these criteria, the following hospitals and health centers were designated as sentinel sites: Mbarara, Fort Portal, and Arua Regional Referral Hospitals; Tororo, Koboko, and Kiryandongo District Hospitals; Kibuli and Nsambya Private Hospitals; and Kiswa and Kitebi Health Center IVs.

These sites are strategically distributed across four major geographical regions of Uganda: Central (Kitebi, Kiswa, Nsambya, Kibuli), Eastern (Tororo), Western (Fort Portal, Mbarara, Kiryandongo), and Northern (Arua, Koboko). Together, these facilities serve a catchment population of approximately 5–7 million people, covering both urban and rural settings (Figure 1).

**Figure 1:**
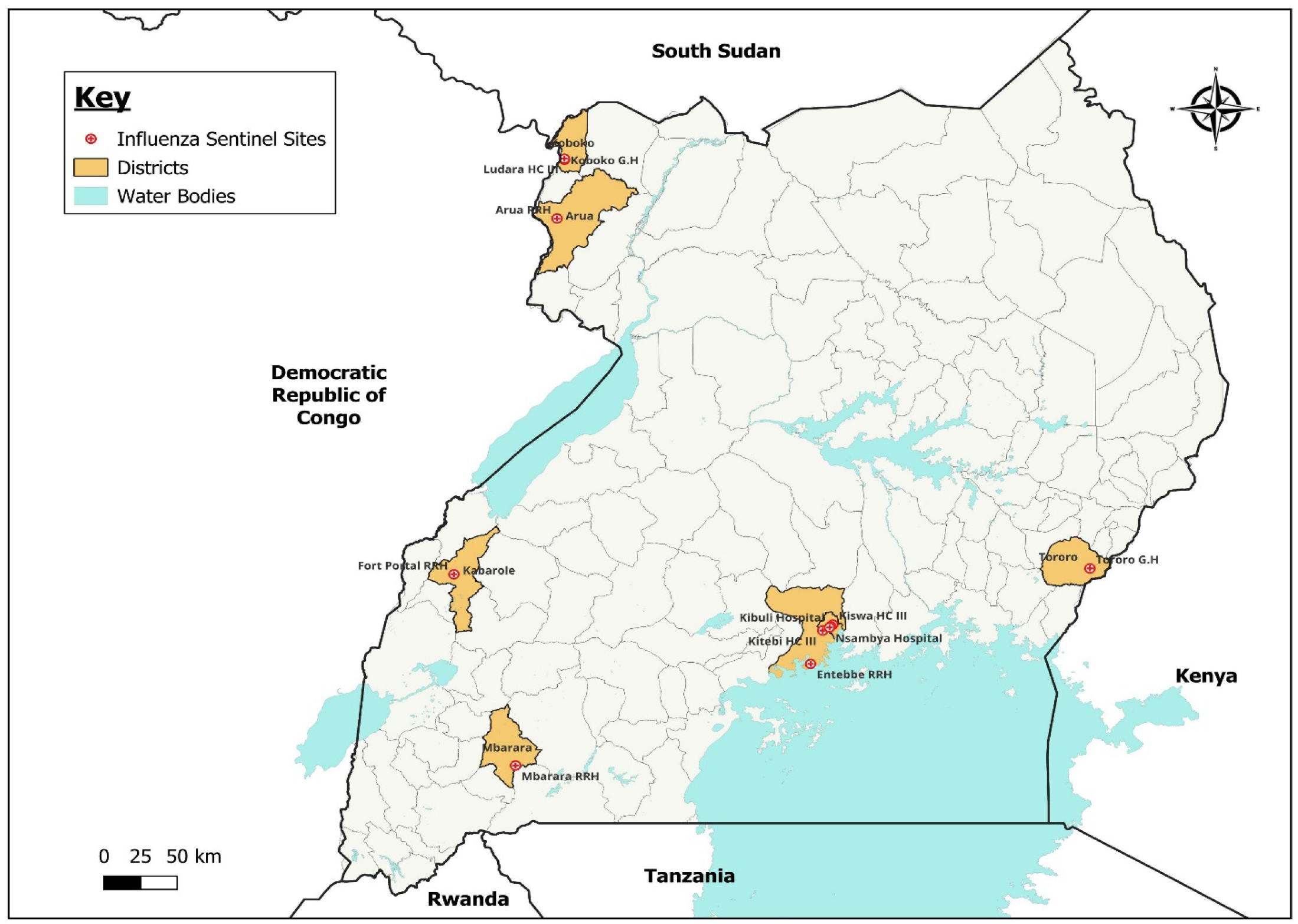
Influenza Sentinel sites in Uganda during 2019-2013.

**Figure 2:**
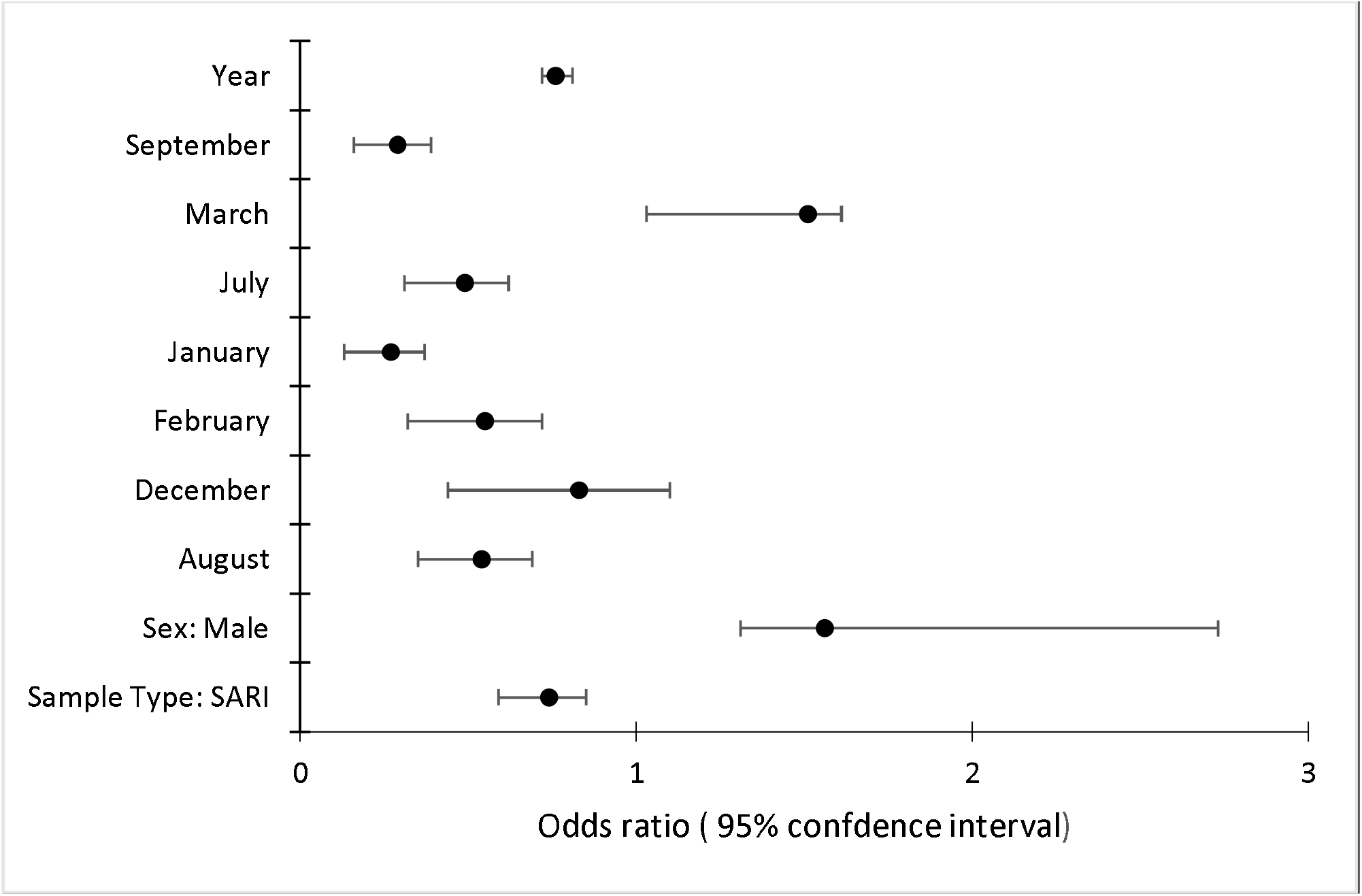
Factors associated with Influenza positivity in Uganda, 2019-2023.

### Participant Case Definitions

This study employed standardized case definitions, adapted from the World Health Organization (WHO), to identify cases of Influenza-Like Illness (ILI) and Severe Acute Respiratory Infection (SARI). ILI was defined as a respiratory infection characterized by a measured fever of ≥38°C, cough, and/or sore throat, with symptom onset within 3 to 10 days prior to presentation, and not requiring hospitalization. SARI was defined as an acute respiratory infection with a history or measurement of fever ≥38°C, cough, shortness of breath, or difficulty breathing, with symptom onset within 10 days of presentation, and requiring hospitalization. These definitions adhered to the WHO ILI/ SARI case definitions and its national adaptation(17).

### Inclusion and Exclusion Criteria

The study population comprised patients presenting at designated influenza sentinel sites between 2019 and 2022, who met the aforementioned ILI or SARI case definitions. Patients were excluded if their respiratory symptoms were attributed to known non-infectious causes, such as asthma exacerbation or pulmonary embolism. Additionally, individuals with incomplete clinical data, those who refused to provide informed consent, or those who did not meet the national guidelines criteria for the respective disease were excluded from the study

### Patient Enrollment, Specimen Processing, and Reporting

At influenza sentinel surveillance sites, clinicians interview eligible patients who meet the standard case definitions for ILI or SARI as outlined above, using a standardized questionnaire to collect demographic, clinical, and epidemiological information. Nasal and or oral pharyngeal swabs are collected and sent to the Uganda Virus Research Institute National Influenza Center, where they undergo Real Time-Polymerase Chain Reaction (RT-PCR) testing for influenza A and B, using primers provided by the United States Centers for Disease Control and Prevention (US CDC). Positive samples for influenza A are further subtyped to identify strains such as A/H1, A/H3, A/H5, and pandemic A/H1N1 (A/H1N1pdm09) using PCR with specific primers and probes (18). The collected data is entered into the National Influenza Centre database for analysis and monitoring. All results are reported weekly to the WHO through the Respmart system and to the Ministry of Health via the National Public Health Emergency Operations Centre. A case investigation form utilized to collect data at the sentinel site is included in the supplementary files.

### Data abstraction, data source, and study variables

We utilized data on patients tested for seasonal influenza during 2019-2023 generated by the national sentinel influenza surveillance system and whose database is stored at the national influenza laboratory at Uganda Virus Research Institute (UVRI). We abstracted data on test result, age, sex, residence, date of sample collection, date of symptom onset and test result.

### Data analysis

Descriptive statistics were used to characterize the data, followed by multivariable logistic regression to identify independent predictors of influenza positivity. The logistic regression model assessed the association of various factors, including sex, sample type, month of the year, and year, with influenza positivity, and provided odds ratios, confidence intervals, and p-values to determine the strength and significance of these associations.

### Ethical considerations

Influenza surveillance data collected between 2019 and 2023 were accessed for research purposes between December 2023 and January 2024. The dataset was de-identified prior to analysis, and the authors did not have access to information that could identify individual participants during or after data collection. The data were obtained from the National Influenza database with permission from the National Influenza Centre. Additionally, the Uganda Public Health Fellowship Program, under which this project was conducted, had permission to access and analyze surveillance data for public health decision-making and dissemination through scientific publications. The U.S. Centers for Disease Control and Prevention (CDC) determined that this study did not constitute human subjects research and was conducted in accordance with applicable laws and policies.

## Results

Of the 18,035 samples tested, 937 (5.2%) were positive for influenza. The positivity rate was higher among patients with Influenza-Like Illness (ILI), with 752 of 13,148 samples (5.7%) testing positive, compared to 185 of 4,895 samples (3.8%) from patients with Severe Acute Respiratory Infection (SARI). Among positive cases, influenza A accounted for the majority (659, 69.7%), while influenza B accounted for 278 cases (29.7%). Of the influenza A positives, A/H3N2 was the predominant subtype (429, 65.1%), followed by A/H1N1pdm09 (221, 33.5%). A small number were unsubtypeable (2, 0.1%) and one case (0.05%) co-infected with A/H3N2 and A/H1N1pdm09, and six cases (0.1%) co-infected with A/H1N1pdm09, A/H3N2, and B/Victoria. All influenza B cases were identified as the B/Victoria lineage.

### Seasonal and Demographic Predictors of Influenza Positivity

Multivariable logistic regression revealed significant associations between several factors and influenza positivity, after adjusting for relevant covariates. Males exhibited a 1.56-fold higher odds of influenza positivity compared to females (OR: 1.56, 95% CI: 1.35–1.81, p < 0.001). Conversely, individuals sampled under the Severe Acute Respiratory Infection (SARI) case definition were significantly less likely to test positive than those sampled under the Influenza-like Illness (ILI) definition (OR: 0.74, 95% CI: 0.61–0.89, p = 0.006).

Seasonal variation in influenza positivity was evident. March was associated with increased odds of positivity (OR: 1.51, 95% CI: 1.16–1.99, p = 0.001), while several other months, notably September and January, showed significantly reduced odds compared to April (reference month) (OR: 0.29, 95% CI: 0.19–0.42, p = 0.001 and OR: 0.27, 95% CI: 0.17–0.41, p = 0.001, respectively). A significant inverse association was observed between year and influenza positivity (OR: 0.76, 95% CI: 0.71–0.80, p < 0.001), indicating a decline in positivity over the study period.

Geographic region did not significantly influence influenza positivity. Odds ratios for the Eastern, Northern, and Western regions, compared to the Central region, were not statistically significant (OR: 1.00, 95% CI: 0.77–1.29, p = 0.977; OR: 1.14, 95% CI: 0.84–1.52, p = 0.398; and OR: 1.11, 95% CI: 0.85–1.43, p = 0.428, respectively)

### Positivity rate of seasonal influenza in Uganda by region, 2019-2023

The positivity rate ranged from 5% in the Central Region to 8% in the Western Region. The Western and Southwestern subregions, including Bunyoro, Rwenzori, Greater Toro, Ankole, and Kigezi, indicated the highest positivity rates of 8% (Figure 3).

**Figure 3:**
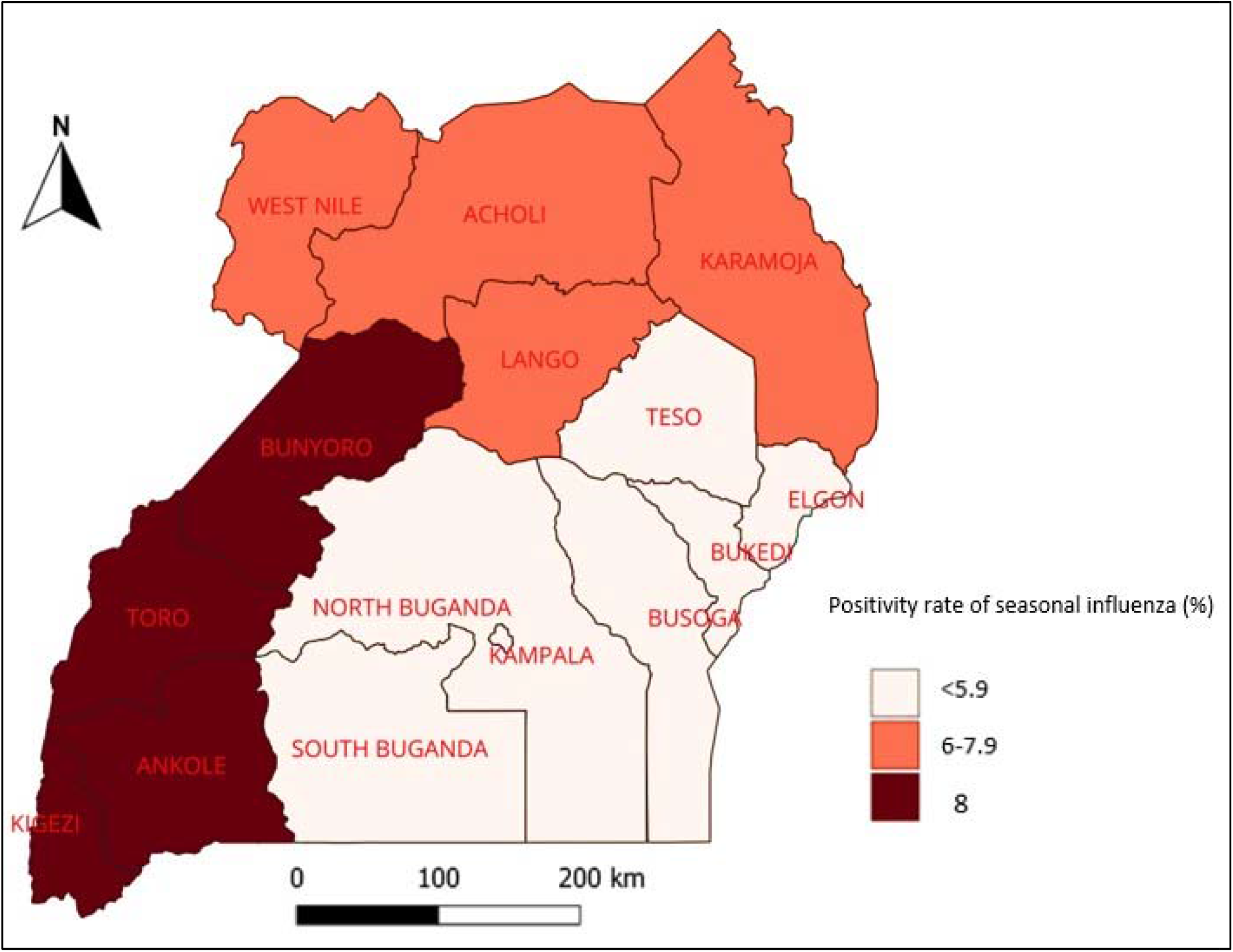
Positivity rate of seasonal influenza in Uganda by region, 2019-2023.

### Seasonal trends in Positivity Rates by year from 2019-2023

The overall positivity rate showed a marked upward trend, rising from 6.8% in 2019 to a peak of 44% in 2022, before declining to 25% in 2023. The number of samples tested increased substantially over the years, from 1,234 in 2019 to a maximum of 7,992 in 2022. The highest number of positive cases was recorded in 2022, accounting for 44% of all tested samples, indicating a possible outbreak or heightened transmission that year. This was followed by a decline in 2023, although the positivity rate (25%) remained higher than pre-2021 levels.

## Discussion

Over the study period, influenza was more commonly detected among patients presenting with Influenza-Like Illness (ILI) than those with Severe Acute Respiratory Infection (SARI). This observation aligns with previous reports suggesting that ILI cases may present earlier in the course of illness, when viral shedding is at its peak, leading to improved detection of influenza viruses. In contrast, SARI cases often represent more advanced stages of illness or may involve co-infections, which can obscure the identification of influenza as the primary cause (19).

The year-round circulation of influenza in Uganda with distinct seasonal peaks reflects patterns observed in other tropical countries, where influenza does not adhere to the conventional winter-season trend seen in temperate zones (20). Instead, influenza activity in these regions is often associated with rainy seasons and fluctuations in humidity and temperature, which create favorable conditions for viral transmission (21).

Seasonal Influenza type A was the predominant strain throughout the study period. This finding is similar to other studies that revealed Influenza A to be predominant in Uganda (22). Influenza type A viruses are highly mutable pathogens that undergo frequent genetic changes through antigenic drift and shift processes, facilitating the emergence of novel strains. These genetic variations allow the virus to evade host immune defenses, contributing to the occurrence of seasonal epidemics and, occasionally, more severe pandemics (23). Among the Influenza A serotypes, H3N2 was the most prevalent, due to its ability to rapidly evolve and adapt (24). This adaptability enhances its transmissibility and enables it to evade immune responses more effectively, making it a dominant driver of seasonal influenza outbreaks. H3N2 has been consistently associated with seasonal influenza activity in Uganda, as reported in other studies (25). This subtype has also been linked to more severe disease, especially in older populations, and poses a challenge for vaccine formulation due to its rapid evolution(26). The persistence of influenza B, particularly the B/Victoria lineage, also emphasizes the importance of including both influenza A and B components in seasonal vaccines.

While geographic differences in influenza positivity were observed, these did not persist after adjusting for other factors. This may reflect improvements in diagnostic capacity, surveillance expansion, or reporting consistency across regions. It also reinforces the need to interpret regional variations cautiously, particularly when based on proportions alone.

Seasonal variation in influenza activity in Uganda appears to align with periods of increased rainfall, particularly during the primary rainy season in the first quarter of the year. While this pattern suggests an influence of environmental factors on influenza transmission, it is important to note that in sub-Saharan Africa, the rainy season does not consistently coincide with cooler temperatures. This complicates the interpretation of seasonality, as the mechanisms driving influenza circulation in tropical regions may differ from those in temperate zones. Although increased humidity and rainfall may contribute to viral persistence and transmission, the exact environmental drivers remain unclear. Therefore, more region-specific studies are needed to clarify the role of climatic factors beyond rainfall alone in shaping influenza seasonality in tropical settings like Uganda.

The highest positivity rates were observed in 2019 and 2023, with notable peaks exceeding 80%. In contrast, 2020 and 2021 showed reduced influenza activity, likely due to disruptions caused by COVID-19-related interventions, such as lockdowns and reduced travel a sharp decrease in the number of cases was also observed in other countries during the COVID-19 outbreak in 2020 and 2021 (28,29).

Males had a higher positivity rate than females, which is consistent with the literature, Jean et al. (30) reported males to have a higher positivity rate for respiratory viruses compared to females (52.8% vs. 50%). Similarly, Rath et al. (31) also observed that among hospitalized patients with influenza, the positivity rate was higher in males than in females (56% vs. 44%). Sex and gender differences significantly influence immune responses, susceptibility to viral infections, and outcomes, shaped by biological, social, and cultural factors. Females generally exhibit stronger innate and adaptive immune responses than males due to factors like sex steroids (e.g., estrogen and progesterone) and genetic influences, including X-chromosome-linked immunity genes resulting in a weaker immune system to resist infection. (32,33). Additionally, behavioral and occupational exposure factors could also play a role, although further research is needed to fully understand these dynamics in the Ugandan context.

The COVID-19 pandemic significantly impacted influenza trends, with health system priorities shifting toward the COVID-19 response, leading to reduced influenza testing and reporting as observed in 2020 and 2021. Additionally, non-pharmaceutical interventions such as mask-wearing, social distancing, and travel restrictions likely contributed to the sharp decline in influenza circulation observed during 2020 and 2021(34).

Finally, while this study provides important insights, caution is warranted in interpreting prevalence differences as causal. Observed associations may be influenced by a range of factors including testing practices, healthcare-seeking behavior, and surveillance quality. Future studies should aim to explore these associations using more robust designs, including cohort or case-control approaches, to establish causality and support targeted interventions.

## Conclusion

The observed seasonal patterns and demographic differences in influenza positivity have important implications for public health planning and response. Recognizing that influenza activity in Uganda aligns with rainy seasons suggests that surveillance, vaccination campaigns, and public awareness efforts could be strategically intensified ahead of these peak periods. The predominance of Influenza A (H3N2) and its association with more severe outcomes highlights the need for timely and strain-specific vaccine formulations. Additionally, the higher positivity observed among males and younger age groups may warrant targeted messaging and interventions aimed at these populations to reduce transmission risk. These findings support the need for strengthening routine surveillance, enhancing diagnostic capacity, and considering the introduction or expansion of seasonal influenza vaccination programs, particularly for high-risk groups. Overall, integrating influenza preparedness into broader seasonal disease control strategies can improve epidemic readiness and reduce disease burden.

## Supporting information

CDC Clearance

Permission to use public Data

## Data Availability

All data produced in the present work are contained in the manuscript

## List of abbreviations

CDC: Centers for Disease Control and Prevention
NIC: National Influenza Centre
PCR: Polymerase Chain Reaction
UVRI: Uganda Virus Research Institute
WHO: World Health Organisation

## Declarations

### Ethics approval and consent to participate

This evaluation was done to inform public health practice and therefore determined as non-research. The Uganda Virus Research Institute, the custodian of the Influenza surveillance data, granted administrative clearance for accessing and using the data in the database for this analysis.

Data held on computers were encrypted with a password which was made available to the data analysis team.

### Consent for publication

Not Applicable

### Availability of data and materials

The datasets upon which our findings are based belong to the Uganda Virus Research Institute. For confidentiality reasons, the datasets are not publicly available.

However, the datasets can be availed upon reasonable request from the corresponding author with permission from the Uganda Virus Research Institute.

### Funding and disclaimer

Not applicable

### Authors’ Contributions

NAM: Participated in the conception, design, analysis, and interpretation of the study and wrote the draft manuscript; THN, GS, NO and GB reviewed the report, reviewed the drafts of the manuscript for intellectual content and made multiple edits to the draft manuscript; JK, JJL, ARA reviewed the manuscript to ensure intellectual content and scientific integrity. All authors read and approved the final manuscript.

## Acknowledgements

The authors thank sentinel site clinicians and the staff of the Uganda Virus Research Institute National Influenza Center for their assistance with data collection and laboratory testing.

