## Supplementary material for "Types, Subtypes and Positivity Rates of Seasonal Influenza in Uganda, 2019–2023": CDC Clearance: Martha_NRD_Desc_Influenza_cleared.docx

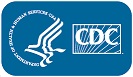
Project Determination

### **Descriptive analysis of National Influenza Center laboratory data in Uganda from 2019-2023**

| **Project ID:** | 0900f3eb8231e873 |
| --- | --- |
| **Accession #:** | CGH-EAFRB-2/29/24-1e873 |
| **Project Contact:** | Gloria Bahizi |
| **Organization:** | CGH/DGHP/EAFRB |
| **Status:** | Project In Progress |
| **Intended Use:** | Project Determination |
| **Estimated Start Date:** | 02/29/24 |
| **Estimated Completion Date:** | 01/01/25 |
| **CDC/ATSDR HRPO/IRB Protocol#:** |  |
| **OMB Control#:** |  |

Description

Priority

Standard

Date Needed

03/08/24

Determination Start Date

02/29/24

Description

Influenza, also known as flu, is a viral infection of the nose, throat, and lungs. The disease is highly contagious, with person-to-person spread by aerosol droplets. Influenza contributes to severe acute respiratory infections, which are the third leading cause of death globally, with 4.25 million deaths annually. The World Health Organization (WHO) estimated the annual mortality burden of influenza to be 250,000 to 500,000 deaths globally. In Africa, data have shown influenza to be a major cause of respiratory illness, especially in children. As with other tropical and sub-tropical countries, influenza viruses circulate in Uganda throughout the year with two peaks: the major one from September to November and a minor one from March to June. Factors for influenza seasonality and spatial distribution are not fully understood but likely reflect individual, population, and environmental determinants. To understand the burden and epidemiology of severe acute respiratory infections and influenza in Uganda, the Uganda Virus Research Institute (UVRI) implemented a national hospital-based sentinel surveillance system to detect and monitor the epidemiology of seasonal influenza and novel influenza viruses as well as other respiratory pathogens of public health interest. Currently, there are 18 sentinel sites located in Kampala, Wakiso, Tororo, Mukono, Mbarara, Kabarole, Arua, Kiryandongo, and Koboko districts. Nasopharyngeal and/or oropharyngeal swab samples, and demographic and clinical data are collected from patients presenting with influenza-like illness (ILI) and those hospitalized with severe acute respiratory illness (SARI) at the sentinel sites. Samples are then routinely shipped frozen to the National Influenza Centre (NIC) laboratory at UVRI for testing. The samples are tested for influenza A and B, Respiratory Syncytial Virus (RSV), and SARS-CoV-2, by real-time reverse transcriptase polymerase chain reaction (rt RT-PCR). All positive influenza A samples are subjected to a second rt RT-PCR test to determine the subtype. This analysis seeks to determine the prevalence, distribution, and patterns of circulating influenza in Uganda from 2019 to 2023. This will inform policymakers on the types and epidemiology of circulating respiratory viruses, for preparedness, prevention, and clinical treatment strategies such as vaccination to prevent outbreaks. In addition, continuous surveillance of influenza and other respiratory viruses in Uganda will enhance its ability to respond effectively to respiratory infections and protect its population.

IMS/CIO/Epi-Aid/Lab-Aid/Chemical Exposure Submission

No

IMS Activation Name

Not selected

Select the primary priority of the project

Not selected

Select the secondary priority(s) of the project

Not selected

Select the task force associated with the response

Not selected

CIO Emergency Response Name

Not selected

Epi-Aid Name

Not selected

Lab-Aid Name

Not selected

Assessment of Chemical Exposure Name

Not selected

Goals/Purpose

This project will describe the prevalence, distribution, and patterns of circulation of influenza in Uganda from 2019 to 2023.

Objective

1. To determine the prevalence of influenza in Uganda from January 2019 to December 2023 2. To describe the distribution of influenza by time, place, and person in Uganda from January 2019 to December 2023

Does your project measure health disparities among populations/groups experiencing social, economic, geographic, and/or environmental disadvantages?

No

Does your project investigate underlying contributors to health inequities among populations/groups experiencing social, economic, geographic, and/or environmental disadvantages?

No

Does your project propose, implement, or evaluate an action to move towards eliminating health inequities?

No

Activities or Tasks

Secondary Data or Specimen Analysis

Target Population to be Included/Represented

International

Tags/Keywords

Influenza, Human; Respiratory Tract Infections; Viruses

CDC's Role

CDC is NOT a recipient or provider of private data, specimens, materials or services; CDC is provider of technical assistance or staff time in the absence of CDC support; CDC is providing funding; CDC provides technical assistance but does not specifically request or approve study design or data collection

Method Categories

Secondary Data Analysis

Methods

This will be a descriptive project using secondary data. We shall abstract retrospective data from the National Influenza Center (NIC) database which is an EPI INFO version 3.5.3 stored at the Uganda Virus Research Institute. This is the national reference laboratory for influenza and COVID-19 in Uganda. The laboratory receives suspected samples from all parts of the country. Data for all samples tested at the National Influenza Center from January 2019 to December 2023 will be included in the study. The study variables will include, dates of sample receipt, test results, age, sex, and the district of residents. Data will be exported to STATA version 13.0 for analysis. We shall calculate the proportions of Influenza cases to determine the trends and prevalence over time and across geographical locations. Data will be presented in graphs, tables, and maps.

Collection of Info, Data, or Bio specimens

Data will be obtained from the National Influenza Centre (NIC) database at the Uganda Virus Research Institute. Data from January 2019 to December 2023 will be extracted as an excel sheet from the database. No personally identifiable information will be used in this project.

Expected Use of Findings/Results and their impact

Results from this study will inform policymakers and the public about the burden of Influenza. Results will be presented to the Ministry of Health and submitted for presentations at national and international conferences to provide evidence for policy action. We plan to publish the results in a peer-reviewed journal and the Uganda National Institute of Public Health Bulletin.

Could Individuals potentially be identified based on Information Collected?

No

| ****Funding**** |  |  |  |  |  |
| --- | --- | --- | --- | --- | --- |
| Funding Type | Funding Title | Funding # | Original Fiscal Year | # of Years of Award | Budget Amount |
| CDC Cooperative Agreement | Strengthen Capacity of Uganda Ministry of Health and Sub - National Entities to Execute Essential Public Health Functions through supporting Public Health Workforce Development | 2356 | 2021 | 5 |  |

****HSC Review****

HSC Attributes

Non-Human/Non-Animal Material

Yes

****Regulation and Policy****

Do you anticipate this project will need IRB review by the CDC IRB, NIOSH IRB, or through reliance on an external IRB?

No

Will you be working with an outside Organization or Institution? Yes

| ****Institutions**** |  |  |  |  |
| --- | --- | --- | --- | --- |
| Institution | FWA # | FWA Exp. Date | Funding | Funding Restriction Amount |
| Makerere University School of Public Health | FWA00011353 | 07/28/28 | Strengthen Capacity of Uganda Ministry of Health and Sub - National Entities to Execute Essential Public Health Functions through supporting Public Health Workforce Development - 2356 |  |

| Institution | Funding Restriction Percentage | Funding Restriction Reason | Funding Restriction has been lifted |
| --- | --- | --- | --- |
| Makerere University School of Public Health |  |  |  |

| Institution | Institution Role(s) | Institution Project Title | Institution Project Tracking # | Prime Institution |
| --- | --- | --- | --- | --- |
| Makerere University School of Public Health | Funding or Sponsoring |  |  |  |

| Institution | Regulatory Coverage | IRB Review Status |
| --- | --- | --- |
| Makerere University School of Public Health | IRB Review is Not Required |  |

| Institution | Registered IRB | IRB Registration Exp. Date | IRB Approval Status |
| --- | --- | --- | --- |
| Makerere University School of Public Health | Makerere U Inst of Public Hlth IRB #1 - HDREC | 02/22/26 |  |

| Institution | IRB Approval Date | IRB Approval Exp. Date | Relying Institution IRB |
| --- | --- | --- | --- |
| Makerere University School of Public Health |  |  |  |

| ****Staff**** |  |  |  |  |  |  |  |  |
| --- | --- | --- | --- | --- | --- | --- | --- | --- |
| Staff Member | SIQT Exp. Date | Citi Biomedical Exp. Date | Citi Social and Behavioral Exp. Date | Citi Good Clinical Exp. Date | Staff Role | Email | Phone # | Organization/  Institution |
| AlexArio | n/a | n/a | n/a | n/a | Co-Investigator | |  |  |
| AnnetNankya | n/a | n/a | n/a | n/a | Principal Investigator | |  |  |
| GloriaBahizi | 07/03/2026 |  |  | 11/09/2026 | Co-Investigator | | - - | EAST AFRICA REGION BRANCH |
| SamuelGiddudu | n/a | n/a | n/a | n/a | Co-Investigator | |  |  |
| Thomas ApolloNsibambi | 10/19/2026 | 02/11/2026 |  |  | Co-Investigator | | - - | UGANDA |

| ****DMP**** |  |
| --- | --- |
| ****Proposed Data Collection Start Date**** | **02/29/24** |
| ****Proposed Data Collection End Date**** | **07/24/24** |
| ****Proposed Public Access Level**** | **Non-Public** |
| ****Reason for not Releasing the Data**** | **Removal of identifiers renders the remaining data of no value** |
| ****Public Access justification**** | **No value** |
| ****How Access Will Be Provided for Data**** | **No value** |
| ****Plans for archival and long-term preservation of the data**** |  |

| ****Spatiality (Geographic Location)**** |  |  |
| --- | --- | --- |
| Country | State/Province | County/Region |
| Republic of Uganda |  |  |

| ****Determinations**** | | | |
| --- | --- | --- | --- |
| Determination | Justification | Completed | Entered By & Role |
| HSC:  Does NOT Require HRPO Review | Research Not Involving Human Subjects  *45 CFR 46.102(e)* | 03/04/24 | Abel_Jason A. (jza5) CIO HSC |
| PRA:  PRA does not apply | Qualifies for a regulatory exclusion: No Information being collected *Justification:* PRA not applicable - Secondary use of existing data only. | 03/04/24 | Abel_Jason A. (jza5) OMB / PRA |
| ICRO:  Concur |  | 03/04/24 | Zirger_Jeffrey (wtj5) ICRO Reviewer |
