## Supplementary material for "Types, Subtypes and Positivity Rates of Seasonal Influenza in Uganda, 2019–2023": Permission to use public Data: MoH Directive-DOC-20221222-WA0009.pdf

General Office: 340874/231563/9
TeleFax 256-41-231584
Telex: 61372 HEALTH UGA.

THE REPUBLIC OF UGANDA

Ministry of Health
P.O. Box 7272
Kampala,
Uganda

In ANY CORRESPONDENCE ON
THIS SUBJECT PLEASE QUOTE
NO. ADM. PCA/198

November 25, 2015

**To: All Stakeholders and Partners**
**All MoH Staff**

**RE: ACCESS AND UTILIZATION OF MINISTRY OF HEALTH DATA AND DATABASE BY PUBLIC HEALTH FELLOWSHIP PROGRAM FELLOWS**

Ministry of Health in collaboration with Makerere University School of Public Health and Centers for Disease Control and Prevention has commenced implementation of the Public Health Fellowship Program as the critical capacity building component of the Uganda National Institute of Public Health. The initial cohort of 10 Field Epidemiology Track Fellows have all been placed in MoH programs and institutions. Every year about a similar number of fellows will be placed in MoH programs and departments. Since the establishment of the program in January 2015, the fellows have made important contributions to the detection, investigation, and control of outbreaks, and collected valuable epidemiologic information to guide public health practices in Uganda, at the same time as they build their competencies in applied epidemiology. As the fellowship program continues to expand its scope, access to and utilization of data are critical not only for the continued success of the fellowship program, but more importantly, so that the data collected by the Ministry of Health are utilized to the fullest extent possible for guiding public health practices in Uganda.

The purpose of this communication is to inform you that the Fellows have been trained in DHIS2 and given access rights and they can comfortably navigate the system to generate necessary reports. More so they are actively involved in outbreak investigations as part of the National Rapid Response Team. They are therefore granted permission to access and analyze MoH data in DHIS2 as well as data generated through surveys or field investigations, so the data can be turned into information for decision-making in the control and prevention of outbreaks, and in other public health programming; additionally, the analysis of these data may also lead to scientific publications as deemed applicable and appropriate by MoH and Public Health Fellowship Programme Secretariat.

Yours Sincerely,

**Dr. Jane Ruth Aceng**
**Director General Health Services**
